# Clinical features and management of severe COVID-19: A retrospective study in Wuxi, Jiangsu Province, China

**DOI:** 10.1101/2020.04.10.20060335

**Authors:** Xiufeng Jiang, Jianxin Tao, Hui Wu, Yixin Wang, Wei Zhao, Min Zhou, Jiehui Huang, Qian You, Meng Hua, Feng Zhu, Xiaoqing Zhang, Meifang Qian, Yuanwang Qiu

## Abstract

**Objective:** We aimed to investigate clinical features and management of 55 COVID-19 patients in Wuxi, especially severe COVID-19.

**Methods:** Epidemiological, demographic, clinical, laboratory, imaging, treatment, and outcome data of patients were collected. Follow-up lasted until April 6, 2020.

**Results:** All 55 patients included 47 (85.5%) non-severe patients and 8 (14.5%) severe patients. Common comorbidities were hypertension and diabetes. Common symptoms were fever, cough and sputum. Lymphopenia was a common laboratory finding, and ground-glass opacity was a common chest CT feature. All patients received antiviral therapy of α-interferon inhalation and lopinavir-ritonavir tablets. Common complications included acute liver injury and respiratory failure. All patients were discharged. No death was occurred and no medical staff got infected. Patients with severe COVID-19 showed significantly older age, decreased lymphocytes, increased C reactive protein, and higher frequency of bilateral lung infiltration compared to non-severe patients. Significantly more treatments including antibiotic therapy and mechanical ventilation, longer hospitalization stay and higher cost were shown on severe patients.

**Conclusions:** Our study suggested that patients with severe COVID-19 may be more likely to have an older age, present with lymphopenia and bilateral lung infiltration, receive multiple treatments and stay longer in hospital.

## Introduction

Since December 2019, a cluster of pneumonia cases of unknown origin that manifested fever, cough and dyspnea have been identified in Wuhan, Hubei Province of China[1]. The pathogen which was soon isolated and identified is a novel enveloped RNA beta-coronavirus and has recently been named as severe acute respiratory syndrome coronavirus 2 (SARS-CoV-2) by the World Health Organization (WHO)[2]. The pneumonia has also been recognized as coronavirus disease 2019 (COVID-19) by WHO and it poses a huge threat to the global world, which has developed into a pandemic[3]. Severe COVID-19 has caused a big burden of the society. By April 6, 2020, over 1,200,000 cases have been identified worldwide, in which a total of over 70,000 patients died.

The outbreak of COVID-19 has been reported to be associated with person-to-person transmission, and a considerable portion of the disease is caused by epidemic clusters, especially familial clusters[4]. Gan et al. collected the data of 1,052 cases in 366 epidemic clusters among 36 cities in China, 86.9% of which occurred in families[5]. Multiple studies have been published to describe the epidemiological and clinical characteristics of COVID-19 in Wuhan. Zhou et al. and Chen et al. reported 191 cases and 99 cases, respectively, indicating that SARS-CoV-2 is more likely to infect the clustered elderly with comorbidities, leading to serious and fatal respiratory failure[6, 7]. However, the features of COVID-19 still need further investigation due to limited reported cases and few studies reported the characteristics of COVID-19 out of Wuhan.

Wuxi, one of the large cities in Jiangsu Province, has totally 55 people infected with SARS-CoV-2 out of 6.5 million population since first case was diagnosed on January 23, 2020. Till March 15, all these 55 patients were successfully cured and no medical staff was infected. Herein, we aimed to delineate clinical features and management of 55 COVID-19 patients in Wuxi, especially severe COVID-19. We also would like to provide data update for COVID-19 and share our experience with health care peers and other people who are interested.

## Methods

### Study design and data source

In this retrospective study, we investigated 55 COVID-19 patients in Wuxi, Jiangsu Province, China. All the patients were diagnosed with COVID-19 according to WHO interim guidance and Chinese management guidelines[8, 9]. This study was approved by the Ethics Commission of Wuxi Fifth People’s Hospital (2020-007-1), a designated infectious diseases hospital of COVID-19 and informed consent from participants was also waived by the Ethics Commission.

### Procedures

Epidemiological, demographic, clinical, laboratory, imaging, treatment, and outcome data were retrieved from electronic medical records of patients. Follow-up for this study lasted until April 6, 2020. If data were missing or vague, we confirmed by direct communication with health care providers. All data were collected by two independent doctors (JXF & WH) and checked by another doctor (TJX).

To identify COVID-19, laboratory confirmation of SARS-CoV-2 was performed in our hospital and Wuxi Center for Disease Control and Prevention (CDC) by testing throat swab specimens from upper respiratory tract using the standard protocol of real-time reverse-transcriptase polymerase chain reaction (RT-PCR) assays[10]. All patients received routine blood examinations including complete blood count, liver and renal function test, coagulation test, cardiac enzyme, electrolytes, C reactive protein, procalcitonin and arterial blood gas analysis. Chest X-ray or CT scan were also done for every patient. The criteria for discharge were body temperature back to normal level for more than 3 days, significant amelioration of respiratory symptoms, obvious improvement of acute lung infiltration in chest radiographs, and two consecutive throat swab or sputum specimens tested negative for SARS-CoV-2 at least 24 hours apart[9].

### Definitions

The severity of COVID-19 was defined according to the Chinese management guideline (7th edition). The severe-type patients were characterized by dyspnea, respiratory frequency ≥30/minute, blood oxygen saturation ≤93%, PaO_2_/FiO_2_ ratio <300, and/or lung infiltrates >50% within 24-48 hours[9, 11]. Clustered epidemic was defined as at least two confirmed cases or asymptomatic infections were found within a small area (e.g. a family, a construction site, a work unit, etc.) within 14 days, and there existed the possibility of person-to-person transmission due to close contact, or the possibility of infection due to co-exposure[12]. Fever was defined as axillary temperature >37.2°C. Diarrhea was diagnosed as the passing of loose stools >3 times per day. Acute respiratory distress syndrome (ARDS) was diagnosed according to the Berlin Definition[13]. Respiratory failure was diagnosed as partial arterial oxygen pressure <60mmHg. Acute kidney injury was defined according to KDIGO guideline[14]. Acute liver injury was diagnosed if was alanine aminotransferase >50U/L or aspartate aminotransferase >40U/L[3]. Acute cardiac injury was diagnosed if serum levels of cardiac biomarkers were above the 99th percentile upper reference limit, or if new abnormalities were shown in electrocardiography and echocardiography[10]. Secondary infection was diagnosed when patients showed clinical symptoms or signs of pneumonia or bacteremia and a positive culture of a new pathogen was isolated from lower respiratory tract specimens or blood samples[10]. Shock was defined according to the 2016 Third International Consensus Definition[6].

### Statistical analysis

Continuous variables were described using median (interquartile range, IQR) and comparisons between groups were performed using Mann-Whitney U test. Categorical variables were presented as number (%) and compared by χ^2^ test or Fisher’s exact test between groups as appropriate. Clinical curves were developed using the Kaplan-Meier method with log-rank test. A two-sided α of less than 0.05 was considered statistically significant. Statistical analyses were done using SPSS (version 26.0) or Graphpad Prism (version 8.0).

## Results

### Epidemiological and demographic features

From January 23 to February 16, 2020, 55 patients infected with SARS-CoV-2 were diagnosed and confirmed in Wuxi, Jiangsu Province according to Wuxi CDC. As of March 30, 2020, no new confirmed cases have appeared since February 17. All patients recovered from COVID-19 and were discharged after treatment by March 15 (Figure S1). Patients were divided into two groups according to severity as non-severe type (47 [85.5%]) and severe type (8 [14.5%]). The median age of all patients was 45.0 years (IQR 27.0-60.0; Table 1), ranging from 7 years to 85 years. Patients with severe disease were significantly older than patients with non-severe disease (59.0 years vs 20.0 years, *p*=0.021). 27 (49.1%) patients were male and 28 (50.9%) patients were female.

**Table 1.** Demographic and clinical features of patients with COVID-19

| Characteristics | All patients<br>(N=55) | Disease severity |  | *p* value |
| --- | --- | --- | --- | --- |
|  |  | Non-severe type<br>(N=47) | Severe type<br>(N=8) |  |
| Age (years) | 45.0 (27.0-60.0) | 20.0 (26.0-57.0) | 59.0<br>(50.0-73.75) | **0.021** |
| Sex |  |  |  | 0.661 |
| Male | 27 (49.1%) | 22 (46.8%) | 5 (62.5%) |  |
| Female | 28 (50.9%) | 25 (53.2%) | 3 (37.5%) |  |
| Exposure history |  |  |  |  |
| Imported from Hubei | 22 (40.0%) | 18 (38.3%) | 4 (50.0%) | 0.815 |
| Cluster | 43 (78.2%) | 37 (78.7%) | 6 (75.0%) | >0.999 |
| Incubation period (days) | 7.0 (3.5-10.0) | 7.0 (3.75-9.75) | 4.0 (3.0-5.0) | 0.613 |
| Comorbidity | 29 (52.7%) | 23 (48.9%) | 6 (75.0%) | 0.326 |
| Hypertension | 17 (30.9%) | 14 (29.8%) | 3 (37.5%) | 0.982 |
| Diabetes | 9 (16.4%) | 5 (10.6%) | 4 (50.0%) | **0.024** |
| Coronary heart disease | 2 (3.6%) | 1 (2.1%) | 1 (12.5%) | 0.669 |
| Cerebrovascular disease | 1 (1.8%) | 0 | 1 (12.5%) | 0.145 |
| Chronic kidney disease | 1 (1.8%) | 0 | 1 (12.5%) | 0.145 |
| Chronic liver disease | 2 (3.6%) | 2 (4.3%) | 0 | >0.999 |
| Malignant tumor | 2 (3.6%) | 2 (4.3%) | 0 | >0.999 |
| Sign and symptoms |  |  |  |  |
| Fever | 47 (85.5%) | 39 (83.0%) | 8 (100.0%) | 0.587 |
| Cough | 34 (61.8%) | 26 (55.3%) | 8 (100.0%) | **0.018** |
| Sputum | 27 (49.1%) | 20 (42.6%) | 7 (87.5%) | **0.049** |
| Fatigue | 22 (40.0%) | 17 (36.2%) | 5 (62.5%) | 0.310 |
| Headache | 10 (18.2%) | 8 (17.0%) | 2 (25.0%) | 0.964 |
| Diarrhea | 18 (32.7%) | 14 (29.8%) | 4 (50.0%) | 0.472 |
| Dyspnea | 11 (20.0%) | 3 (6.4%) | 8 (100.0%) | **<0.001** |
| Hemoptysis | 2 (3.6%) | 1 (2.1%) | 1 (12.5%) | 0.669 |
| Chest distress | 6 (10.9%) | 5 (10.6%) | 1 (12.5%) | 0.876 |
| Inappetence | 3 (5.5%) | 2 (4.3%) | 1 (12.5%) | 0.915 |
| Nausea or vomiting | 4 (7.3%) | 4 (8.5%) | 0 | >0.999 |

As shown in Table 1, 22 (40.0%) cases were imported from Hubei, most of which were from Wuhan due to long-term living in or short-term traveling to Wuhan. There existed 43 (78.2%) cases in 11 epidemic clusters among patients, averaging approximately 4 cases per cluster, mainly through familial clusters and dining clusters. The median incubation period was 7.0 days (IQR 3.5-10.0). The exposure history and incubation period were similar between patients with non-severe and severe illness.

Here, we described two epidemic clusters – one was an imported familial cluster and the other one was a combination of familial, transportation and dining clusters (Figure 1). Patient 1-6 lived together in Wuhan. Patient 1-4 and patient 5-6 returned to Wuxi on January 22 and January 19, 2020, respectively. They got diagnosed and admitted to hospital since January 24, and Patient 6 got severe disease (Figure 1A). Any of these six patients could be the first one to get infected and in turn transmitted the virus to the other family members. Patient 7-10 took the same fight back to Wuxi from Japan on January 24 and patient 7-9 were from the same family. On the next day, patient 7-8 had dinner with patient 11-15. They got diagnosis and admission since January 30 and patient 7&11 got severe disease (Figure 1B). Patient 7 & 11 were diagnosed as the severe type.

**Figure 1.**
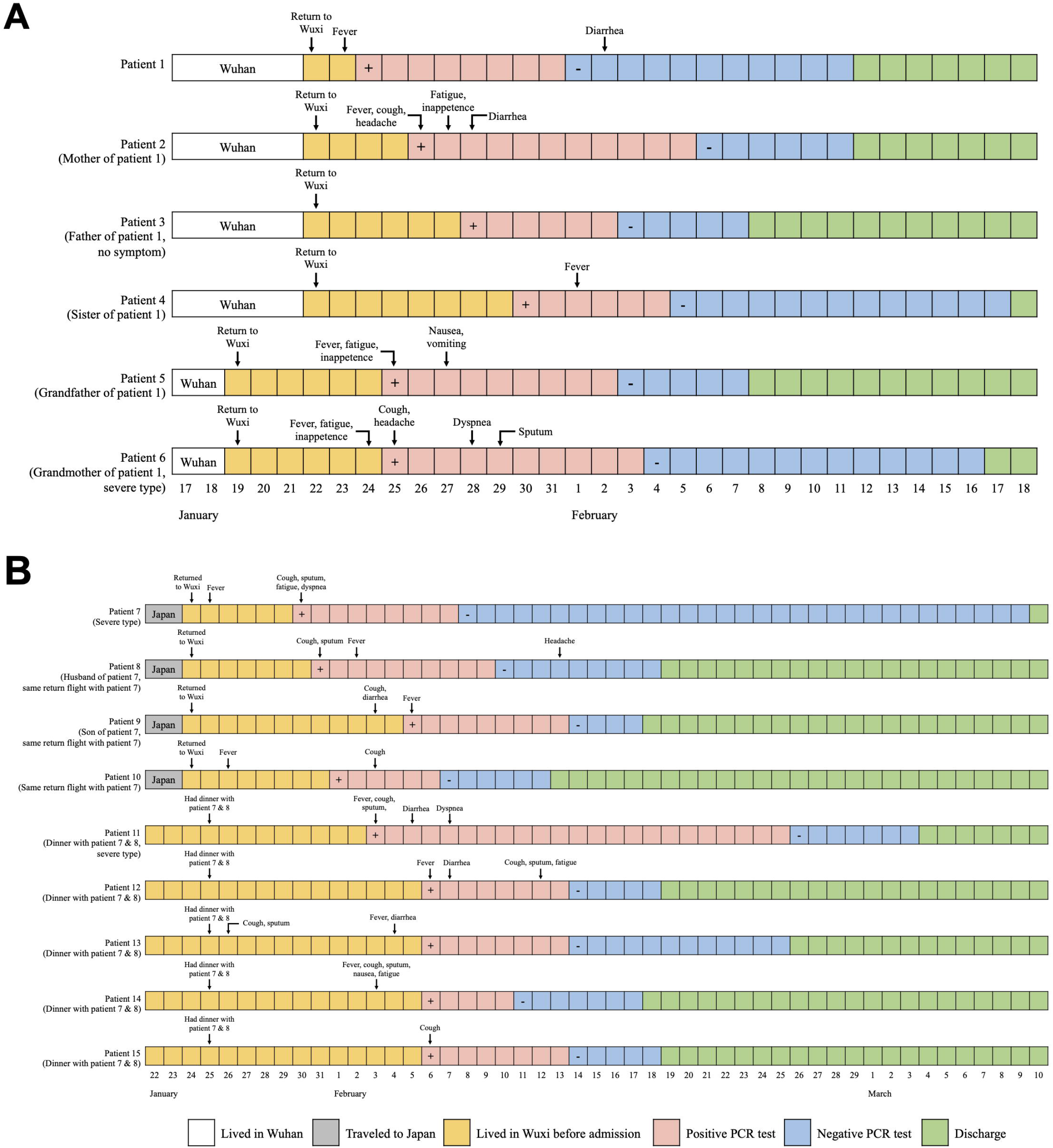
Time course of exposure history, symptoms, laboratory PCR test and discharge in epidemic clusters of COVID-19. (A) An imported familial cluster of six patients. (B) A combination of familial, transportation and dining clusters among nine patients.

### Clinical features

Twenty-nine (52.7%) patients had comorbidities, in which hypertension (17 [30.9%]) was the most common one, followed by diabetes (9 [16.4%]), coronary heart disease (2 [3.6%]), chronic liver disease (2 [3.6%]) and malignant tumor (2 [3.6%]; Table 1). The presence of diabetes was significantly higher in patients with severe disease compared to those with non-severe disease (50.0% vs 10.6%, *p*=0.024). The most common symptoms were fever (47 [85.5%]), cough (34 [61.8%]), and sputum production (27 [49.1%]), followed by fatigue (22 [40.0%]), diarrhea (18 [32.7%]), dyspnea (11 [20.0%]), and headache (10 [18.2%]; Table 1). Patients with severe disease had significantly higher frequency of cough (100.0% vs 55.3%, *p*=0.018), sputum production (87.5% vs 42.6%, *p*=0.049) and dyspnea (100.0% vs 6.4%, *p*<0.001) than those with non-severe disease.

On admission, patients with severe disease significantly lower lymphocytes (0.83×10^9^/L vs 1.40×10^9^/L, *p*=0.010), higher C reactive protein (70.27mg/L vs 4.50mg/L, *p*=0.001) and procalcitonin (0.2ng/mL vs 0.2ng/mL, *p*=0.039) than those with non-severe disease (Table 2). Meanwhile, significantly more patients with severe disease showed increased neutrophils (25.0% vs 0, *p*=0.019) and D-dimer (62.5% vs 21.3%, *p*=0.047) compared to those with non-severe disease. Furthermore, patients also presented decrease in hemoglobin (21 [38.2%]) and albumin (18 [32.7%]), as well as increase in alanine aminotransferase (14 [25.5%]) and aspartate aminotransferase (11 [20.0%]). Thirty-eight (69.1%) patients showed bilateral lung infiltration on chest CT findings, of which patients with severe illness had significantly higher frequency (100.0% vs 63.8%, *p*=0.048) and nine (16.4%) patients showed unilateral lung infiltration (Table 2). Concurrently, ground-glass opacity was presented on 42 (76.4%) patients. The chest CT features of patients with non-severe and severe disease were shown in Figure 2.

**Table 2.**
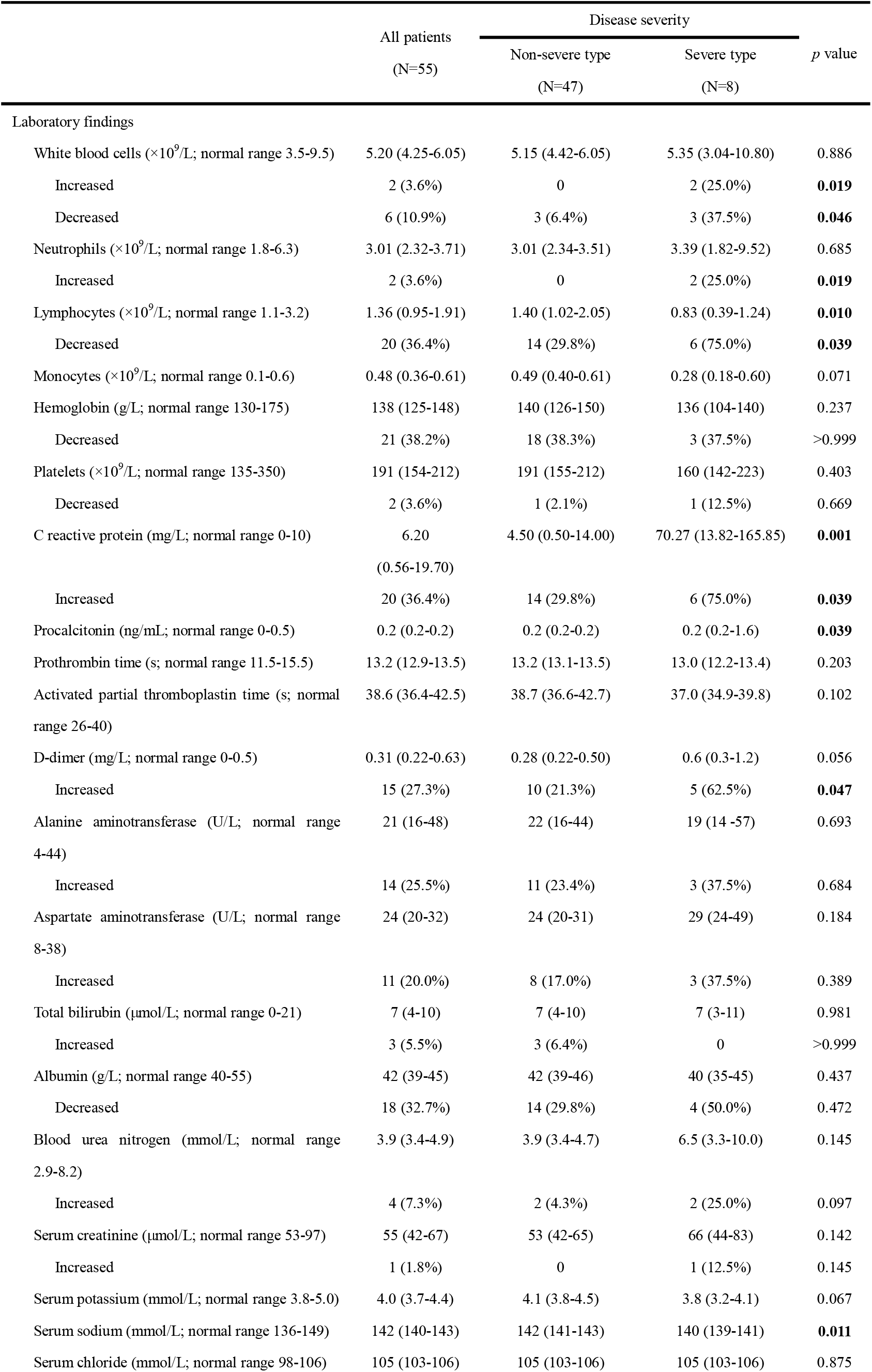
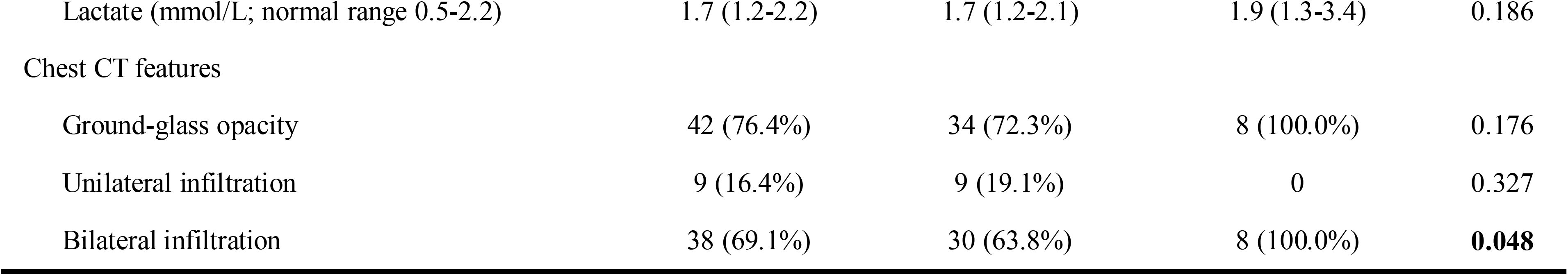
Laboratory and radiological findings of patients with COVID-19

**Figure 2.**
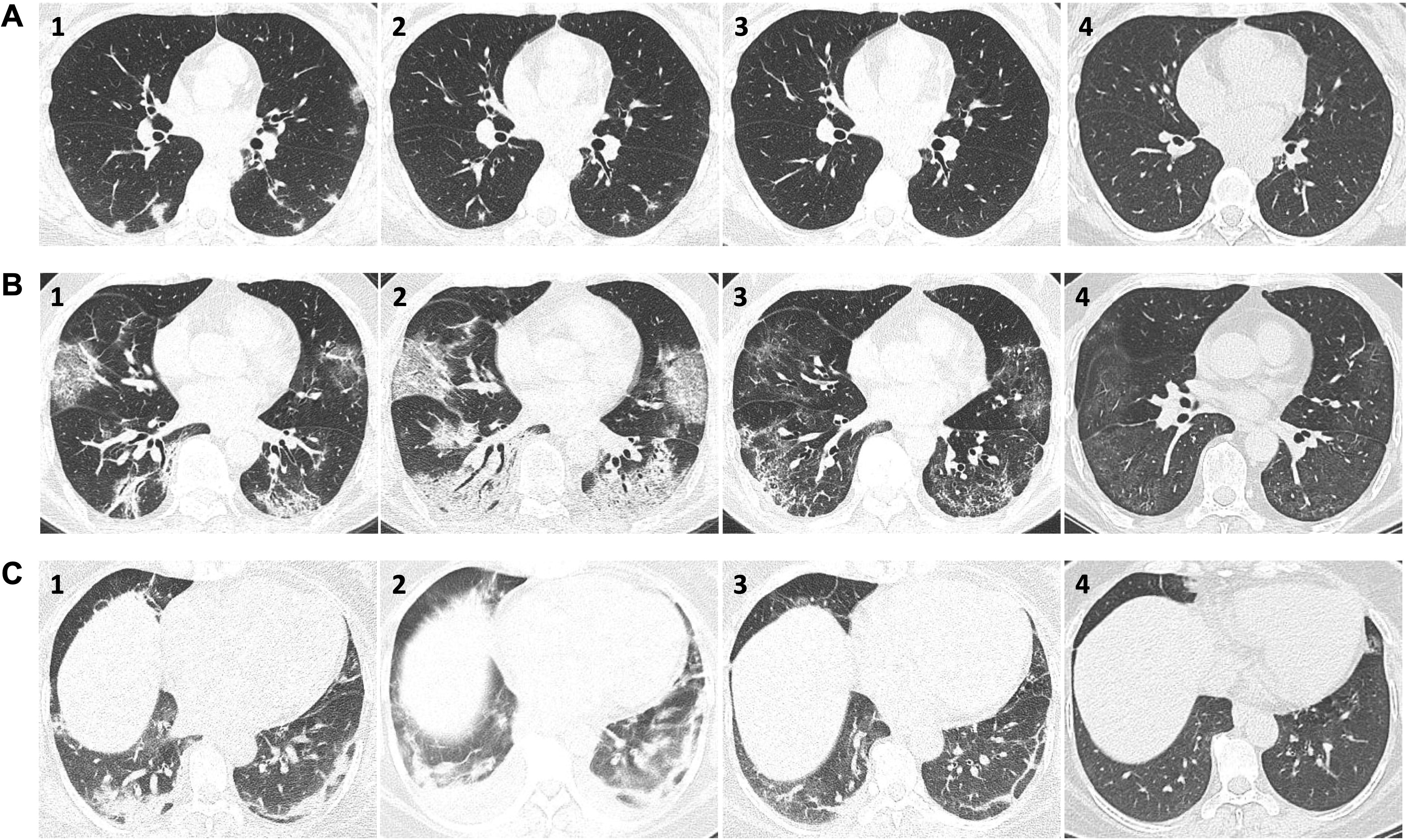
Chest CT features of patients with COVID-19. (A) Chest CT images of 40-year-old woman with non-severe COVID-19. The scan showed multiple scattered patches and nodular ground-glass opacities in bilateral lung filed on admission (A1). Most lesions were absorbed after treatment (A2), and lesions showed completely absorption on the first and second return visit (A3 & 4). (B) Chest CT images of 53-year-old man with severe COVID-19. The scan showed patchy ground-glass opacities with irregular line shadows in bilateral lung filed on admission (B1). Two days later, lesions got deteriorated with increased extent (B2). After treatment, lesions were largely absorbed and turned smaller (B3). On the first return visit, only slight opacities could be observed on both lungs (B4). (C) Chest CT images of 49-year-old woman with severe COVID-19. The scan showed early ground-glass opacities and consolidation in bilateral lower lobes on admission (C1). Three days later, lesions got worsen with increased extent and bilateral pleural effusion (C2). Lesions and pleural effusion were mostly absorbed after treatment (C3) and got completely absorption on the first return visit (C4).

### Treatment and outcomes

All patients received antiviral therapy of α-interferon inhalation and lopinavir-ritonavir tablets (Table 3). 22 (40.0%) patients received arbidol tablets and 4 (7.3%) patients received chloroquine tablets. Meanwhile, many patients received antibiotic therapy (29 [52.7%]), thymosin injection (20 [36.4%]), probiotics tablets (26 [47.3%]) and nasal cannula support (26 [47.3%]). Compared to patients with non-severe disease, patients with severe disease had significantly more frequency to receive chloroquine tablets (37.5% vs 2.1%, *p*=0.005), antibiotic therapy (100.0% vs 44.7%, *p*=0.005), intravenous systemic corticosteroid (75.0% vs 0, *p*<0.001), intravenous immune globulin (50.0% vs 2.1%, *p*<0.001), thymosin injection (100.0% vs 25.5%, *p*<0.001), probiotics tablets (87.5% vs 40.4%, *p*=0.037), low molecular weight heparin (LMWH) injection (75.0% vs 4.3%, *p*<0.001), nasal cannula support (100.0% vs 38.3%, *p*=0.001), high-flow oxygen nasal cannula support (37.5% vs 0, *p*=0.002) and mechanical ventilation (87.5% vs 0, *p*<0.001). Additionally, antifungal therapy, transfusion of convalescent plasma, extracorporeal membrane oxygenation (ECMO) and lung transplant were used on one patient with critically severe disease. The most common complications were acute liver injury (16 [29.1%]) and respiratory failure (10 [18.2%]), followed by ARDS (4 [7.3%]), secondary infection (4 [7.3%]) and acute kidney injury (3 [5.5%]; Table 3). Significantly more patients with severe disease had respiratory failure than those with non-severe disease (100.0% vs 4.3%, *p*<0.001).

**Table 3.** Treatment and outcome of patients with COVID-19

|  | All patients<br>(N=55) | Disease severity |  | *p*<br>value |
| --- | --- | --- | --- | --- |
|  |  | Non-severe type<br>(N=47) | Severe type<br>(N=8) |  |
| Treatment |  |  |  |  |
| Antiviral therapy | 55 (100.0%) | 47 (100.0%) | 8 (100.0%) | -- |
| α-interferon | 55 (100.0%) | 47 (100.0%) | 8 (100.0%) | -- |
| Lopinavir-ritonavir | 55 (100.0%) | 47 (100.0%) | 8 (100.0%) | -- |
| Arbidol | 22 (40.0%) | 19 (40.4%) | 3 (37.5%) | >0.999 |
| Chloroquine | 4 (7.3%) | 1 (2.1%) | 3 (37.5%) | **0.005** |
| Antibiotic therapy | 29 (52.7%) | 21 (44.7%) | 8 (100.0%) | **0.005** |
| Antifungal therapy | 1 (1.8%) | 0 | 1 (12.5%) | 0.145 |
| Corticosteroid | 6 (10.9%) | 0 | 6 (75.0%) | **<0.001** |
| Immune globulin | 5 (9.1%) | 1 (2.1%) | 4 (50.0%) | **<0.001** |
| Thymosin | 20 (36.4%) | 12 (25.5%) | 8 (100.0%) | **<0.001** |
| Probiotics | 26 (47.3%) | 19 (40.4%) | 7 (87.5%) | **0.037** |
| Low molecular weight heparin | 8 (14.5%) | 2 (4.3%) | 6 (75.0%) | **<0.001** |
| Nasal cannula | 26 (47.3%) | 18 (38.3%) | 8 (100.0%) | **0.001** |
| High-flow oxygen nasal cannula | 3 (5.5%) | 0 | 3 (37.5%) | **0.002** |
| Mechanical ventilation | 7 (12.7%) | 0 | 7 (87.5%) | **<0.001** |
| Transfusion of convalescent plasma | 1 (1.8%) | 0 | 1 (12.5%) | 0.145 |
| Extracorporeal membrane oxygenation | 1 (1.8%) | 0 | 1 (12.5%) | 0.145 |
| Lung transplant | 1 (1.8%) | 0 | 1 (12.5%) | 0.145 |
| Complications |  |  |  |  |
| Acute respiratory distress syndrome | 4 (7.3%) | 2 (4.3%) | 2 (25.0%) | 0.176 |
| Respiratory failure | 10 (18.2%) | 2 (4.3%) | 8 (100.0%) | **<0.001** |
| Acute kidney injury | 3 (5.5%) | 1 (2.1%) | 2 (25.0%) | 0.073 |
| Acute liver injury | 16 (29.1%) | 12 (25.5%) | 4 (50.0%) | 0.323 |
| Acute cardiac injury | 1 (1.8%) | 0 | 1 (12.5%) | 0.145 |
| Secondary infection | 4 (7.3%) | 2 (4.3%) | 2 (25.0%) | 0.176 |
| Shock | 1 (1.8%) | 0 | 1 (12.5%) | 0.145 |
| Prognosis |  |  |  |  |
| Discharge | 55 (100.0%) | 47 (100.0%) | 8 (100.0%) | -- |
| Death | 0 | 0 | 0 | -- |
| Duration of hospitalization (days) | 16.0 (13.0-21.0) | 16.0 (13.0-19.0) | 23.0 (17.3-36.3) | **0.003** |
| Duration from admission to negative laboratory test (days) | 8.0 (7.0-11.0) | 8.0 (7.0-11.0) | 9.5 (5.8-18.3) | 0.597 |
| Duration from negative laboratory test to discharge (days) | 7.0 (5.0-10.0) | 6.0 (5.0-9.0) | 14.0 (7.8-19.8) | **0.002** |
| Hospitalization cost (Chinese *yuan*) | 11472.67<br>(8220.14-17509.40) | 10589.70<br>(8184.29-14139.79) | 71902.29<br>(36078.47-97484.76) | **<0.001** |

All patients were discharged from hospital and no death occurred. The median duration of hospitalization among all patients was 16.0 days (IQR 5.0-10.0; Table 3) and patients with severe disease had longer hospitalization compared with those with non-severe disease (23.0 days vs 16.0 days, *p*=0.003; HR=0.37 [95% CI 0.21-0.65], *p*=0.0012; Figure 3A). Patients with severe disease also stayed significantly longer in hospital after negative PCR test (14.0 days vs 6.0 days, *p*=0.002; HR=0.38 [95% CI 0.21-0.66], *p*=0.0010; Figure 3C). The median hospitalization cost was 11472.67 *yuan* (IQR 8220.14-17509.40), in which patients with severe disease spent significantly more money than those with non-severe disease (71902.29 *yuan* vs 10589.70 *yuan, p*<0.001).

**Figure 3.**
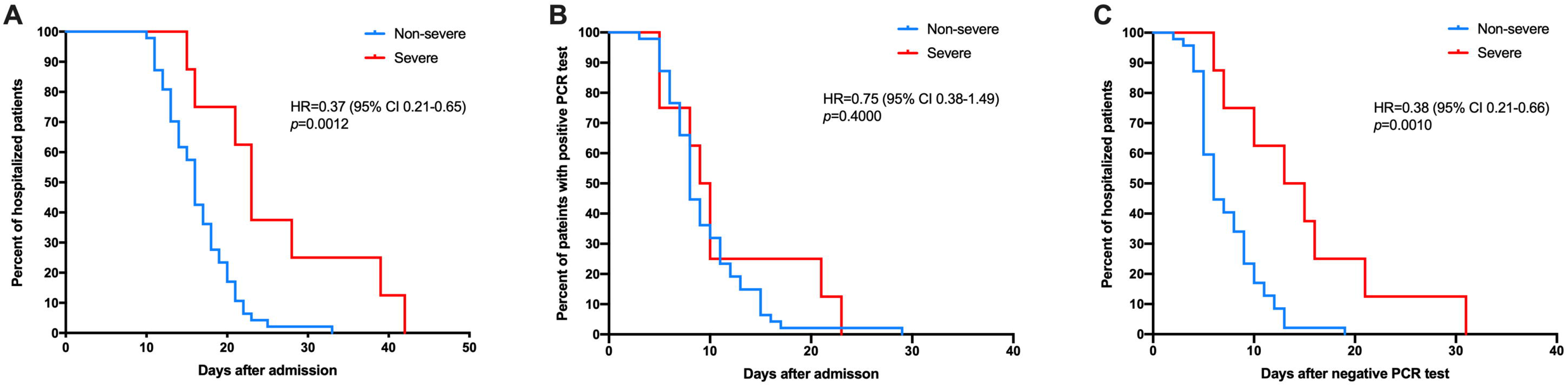
Kaplan-Meier curves of duration of overall hospitalization (A), conversion from positive to negative PCR test (B), and hospitalization after negative PCR test (C) between patients with non-severe and severe COVID-19.

## Discussion

To our knowledge, this report is the first retrospective study of all 55 COVID-19 patients in Wuxi, Jiangsu Province, China. We described epidemiological, demographic, clinical, imaging features as well as treatment and outcomes of all patients. All patients got recovery and discharged from hospital. No death was occurred and no medical staff got infected.

In our study, the median age of patients with severe disease were significantly higher compared to patients with non-severe disease, suggesting that older patients may be more susceptible of getting severe disease than younger patients. However, no significant sex difference was found between patients with non-severe and severe illness. A majority (78.2%) of patients got infected due to epidemic clusters, mostly via familial and dining clusters, which provided the evidence of person-to-person transmission of SARS-CoV-2. According to cluster investigation technical guidelines from the Chinese Epidemiology Working Group, epidemic clusters are divided into several subtypes, which refer to the cluster of family, dining, work unit, transportation and public place[12].

Over half (52.7%) of patients had comorbidities, most of which were hypertension (30.9%) and diabetes (16.4%), whereas we didn’t discover a significant difference of all comorbidities between patients with non-severe and severe disease other than diabetes. Similarly, recent studies showed that the prevalence of diabetes was averagely about two folds higher in severe cases[15]. Common symptoms found in our study were fever, cough, sputum production, dyspnea, fatigue, chest distress and headache. Interestingly, we found 18 (32.7%) patients had diarrhea, with a higher incidence than that in previous studies[11, 16]. Four (7.3%) patients had no signs and symptoms during the onset and they only got the non-severe disease (data not shown). The existence of asymptomatic infected individuals has been reported and they also had the ability of virus transmission to others[17]. Particularly, patients with asymptomatic infection may not realize their illness and they could be important hidden sources of infection, which may bring potential danger to public health. Therefore, it is of great significance for suspected patients to fully cooperate with self-isolation, supervision and screenings.

In laboratory findings, patients with severe disease had significantly lower levels of lymphocytes and decreased serum sodium concentration as well as higher levels of C reactive protein and procalcitonin than those with non-severe disease, implying these abnormalities could be a potential indicator for severe-type COVID-19. However, unlike some other studies, we didn’t find any major differences of leukocytes, neutrophils, D-dimer and liver enzymes between two groups[16, 18]. Our findings also suggested patients with severe disease were more likely to present with increased neutrophils and D-dimer. The results of Chest CT showed a majority of patients had bilateral infiltration and ground-glass opacity, and both of them were presented in all patients with severe disease. Moreover, a small portion of patients showed no obvious abnormalities on CT.

Currently, most of the treatments for COVID-19 are supportive therapy. Till now, no specific drugs or vaccinations have been approved for treatment or prevention from infection with SARS-CoV-2. In our study, all patients received the combination therapy of α-interferon and lopinavir-ritonavir. α-interferon is responsible for broad antiviral effect and enhancement of immune response[19]. Lopinavir, a human immunodeficiency virus type 1 aspartate protease inhibitor, was previously approved for treating SARS-CoV infection in 2003, and ritonavir is combined with lopinavir to increase its plasma half-life through the inhibition of cytochrome P450[20]. Surprisingly, a recent small-scale clinical trial showed no clinical improvement was observed with lopinavir-ritonavir treatment beyond standard care for treating patients with severe disease[20]. However, Deng et al. found that lopinavir-ritonavir combined with arbidol, another drug against SARS-CoV, might show benefit in delaying clinical progression of COVID-19 and attenuating viral transmissibility[21]. Chloroquine is newly recognized as potential effect against SARS-CoV. In our study, chloroquine was used to treat four (7.3%) patients, mostly used on patients with severe disease. Wang et al. discovered chloroquine blocked virus infection in vitro with EC_50_□ of 1.13μM[22] and Yao et al. found hydroxychloroquine was more potent (EC_50_=0.72μM) than chloroquine to inhibit SARS-CoV-2[23], providing more options for drug treatment of COVID-19. Recently, it was reported that remdesivir might have a promising effect against SARS-CoV-2. Remdesivir (GS-5734) is an adenosine analogue, which incorporates into viral RNA chains, and has been confirmed to inhibit SARS-CoV-2 efficiently in vitro with EC_50_□ of 0.77μM[22]. The first patient of COVID-19 in United States showed a good clinical response to remdesivir[24]. Meanwhile, several clinical trials (NCT04257656, NCT04252664 and NCT04292899) are under completion to evaluate the safety and antiviral activity of remdesivir. Overall, it needs to take a long time to identify or invent effective medicine for COVID-19.

From our data, patients with severe disease were more likely to be treated by antibiotic therapy, intravenous systemic corticosteroid, intravenous immune globulin, thymosin injection, probiotics tablets, LMWH injection, high-flow oxygen nasal cannula support and mechanical ventilation, which, to some extent, may explain the longer duration and higher cost of hospitalization in severe patients. Our team also used new techniques to treat the critically severe patient, such as transfusion of convalescent plasma, ECMO, and even lung transplant. No significant difference was found in the duration of conversion from the positive PCR test to the negative one after admission (Table 3 & Figure 3B), however, patients with severe disease had the tendency to stay longer in hospital after the negative PCR test.

SARS-CoV-2, the coronavirus causing COVID-19, has been proved to bind the entry receptor of host cells called angiotensin-converting enzyme II (ACE2), the same as previous SARS-CoV and MERS-CoV[25]. ACE2 is a protein with 805 amino acids and diffusely expressed in heart, vessels, lung, kidney and gut. It functions as the conversion of angiotensin I to angiotensin II, playing a pivotal role in renin-angiotensin-aldosterone system (RAAS)[26]. When infected with SARS-CoV-2, the virus predominantly targeted ACE2 in human alveolar type 2cells, resulting in pulmonary symptoms[27]. It has been reported that ACE2 was associated with comorbidities, symptoms and complications in COVID-19 patients, such as hypertension, diabetes, cardiovascular disease and gastrointestinal abnormalities[28]. However, the mechanism still remains unclear. Chen et al. found that SARS-CoV-2 might attack pericytes of human heart, thus causing capillary endothelial cell dysfunction, leading to cardiovascular disease and cardiac injury[29]. Jin et al. suggested that gastrointestinal epithelial cells had a high affinity to ACE2, indicating the potential of fecal-oral transmission and the tendency of gastrointestinal symptoms in dissemination[3]. Despite SARS-CoV-2 had a huge relationship with ACE2, whether ACE2 could be a therapeutic target still remains controversial and needs further research.

In Wuxi, potent and strict measures have been taken since the outbreak of COVID-19. People need to wear face masks and use Wuxi Health Code when going out. People who have been to Wuhan recently or have had close contact with them would be advised to quarantine at home for at least 14 days. Suspected patients would receive the nucleic acid PCR test and those who got a positive result would be admitted to hospital immediately. Every confirmed patient with COVID-19 would be arranged in the negative pressure ward of a designated infectious hospital for treatment. Those who got discharged still need the PCR test again two and four weeks after discharge, respectively. Of all patients, one patient retested positive of stool samples.

Several limitations do exist in our study. First, this study included only 55 cases thanks to good monitoring and screening in Wuxi, which may cause some statistical and selective bias of our results.

Second, no immune function test like serum T cells or cytokines and some other tests like serum lactate dehydrogenase were involved in the laboratory test. As such their role might be underestimated in causing severe disease. Third, this study was limited to the local investigation, and the findings may be inconsistent with nationwide or worldwide studies, which may decrease its credibility.

## Conclusion

In summary, our retrospective study established the epidemiological and clinical features of all 55 COVID-19 patients in Wuxi, Jiangsu Province, China, and shared some successful experience of treatment, especially patients with severe disease. Our results supported person-to-person transmission of SARS-CoV-2 and suggested that patients with severe COVID-19 may be more likely to have an older age, present with lymphopenia and bilateral lung infiltration, receive multiple treatments and stay longer in hospital. Further large-sized and randomized clinical trials are urgently needed to identify the effective treatments for COVID-19.

## Data Availability

The data used to support the findings of this study are available from the corresponding author upon request.

## Acknowledgement

This study was supported by Wuxi Municipal Health Commission Major Project (Z201811); Wuxi Science and Technology Bureau COVID-19 special project (N2020X009); Wuxi Science and Technology Bureau guiding plan (201812). We would like to thank all medical staff with fighting COVID-19 in Wuxi for their huge contributions.

---

Figure S1. Trend diagram among 55 confirmed cases of COVID-19 in Wuxi, Jiangsu Province, China.

